# The development and usability testing of two arts-based knowledge translation tools for pediatric fractures

**DOI:** 10.64898/2026.07.28.26357986

**Authors:** Shannon D. Scott, Hannah M. Brooks, Samina Ali, Manisha Bharadia, Lisa Hartling

## Abstract

Fractures, especially simple fractures, are a common childhood injury. The purpose of this research was to work with parents to develop and test the usability of an animated video and an interactive infographic about simple fractures in children. Prototypes were designed collaboratively between researchers, healthcare experts, and parents. After refinement, prototypes were evaluated by parents through usability testing in an urban emergency department waiting room in Alberta. Results were positive and overall, the tools were highly rated, suggesting that arts-based digital knowledge translation tools are useful mediums for sharing health information about simple fractures with parents.

## Introduction

Fractures are a common pediatric injury with up to 64% of children sustaining a fracture before the age of 15 years [1]. Among reasons to present to the emergency department (ED), one study in the United States found that fractures accounted for 15% of pediatric visits to the ED [2]. Simple fractures are a type of fracture that is common in pediatric populations and can be treated without surgery. Following discharge from the ED, children often experience significant pain, particularly in the first 3 days following the fracture. As a child recovers, the impact of the injury on a child’s day-to-day life extends to family members, with 40% of caregivers likely to miss work to care for their child following a fracture [3]. Returning to activities without any restrictions takes time and therefore fractures have an impact on a child’s quality of life as they recover.

Qualitative studies [4, 5] exploring the impact of fractures on young children and adolescents’ function found that the areas most significantly impacted in daily life following a fracture were sleep, activities of daily living (e.g. showering, eating), and play (e.g. sports, activities). These reported functional limitations led to emotional struggles, loss of independence, and parental adaptive strategies such as planning, changes to household routines, and missed work [4, 5].

Understanding the experiences and information needs of children and caregivers is essential to creating resources that can effectively communicate health information to help parents make informed decisions when seeking health care for their child who may have a fracture. Furthermore, qualitative studies demonstrate that more effective communication is needed prior to and following discharge to satisfy information needs and guide expectations through the recovery process. Aligning clinician priorities with family experiences can help provide more effective discharge counselling.

Research exploring the benefits of art and narrative based forms of knowledge translation (KT) tools have illustrated the power these forms may have in communicating, engaging, and influencing individuals. Currently, there have been limited numbers of KT tools developed to provide parents education on fractures and how to care for their child following a fracture. The majority of the information currently available is in a strictly written format, or is meant to be used along with an education session by a health care provider (HCP).

The purpose of this research was to develop and assess the usability of an animated video and interactive infographic for families about pediatric fractures. Following prototype completion and usability testing, the finalized tools were made publicly available on our website (echokt.ca) and disseminated through stakeholder websites and on social media.

## Methods

A multi-method study involving patient engagement was used to develop, refine, and evaluate an animated video and interactive infographic for pediatric fractures [6]. Research ethics approval was obtained from the University of Alberta Health Research Ethics Board (Edmonton, AB) [Pro00098397]. Operational approval was obtained from the Stollery Children’s Hospital to conduct usability testing.

### Compilation of Parents’ Narratives

Semi-structured qualitative interviews were conducted by the Pediatric Emergency Advancing Knowledge (PEAK) research team to inform parental and adolescent narratives for the tools. Parents with children who presented to the Stollery Children’s Hospital Emergency Department with long bone fractures that did not require surgical interventions were recruited for qualitative interviews. Parents were asked to share their experiences having a child with the condition, while adolescents were asked to share their experiences having a fracture. Results from the qualitative studies are published elsewhere [4, 5].

### Prototype (Intervention) Development

Results from both the qualitative studies and Bottom Line Recommendations (BLR) developed by Translating Emergency Knowledge for Kids (TREKK) [7] were used to inform the development of an infographic skeleton and video script. The BLR provides evidence-based information for health care professionals (HCPs) including key facts and recommendations for diagnosing and treating fractures in the ED. Following the completion of the infographic skeleton and script, researchers worked with illustrators and graphic designers to develop the tool prototypes. Iterative processes were used to develop the tool prototypes whereby parents, HCPs, and researchers provided several rounds of feedback. HCPs were asked to comment on the clinical relevancy, quality of information and evidence. Parents from our Pediatric Parent Advisory Group (P-PAG) were asked to provide feedback on the length, stylistic elements, and information addressed in the tools.

### Video

The English-language video was 5 minutes and 22 seconds long, narrated in the third person, and included closed captioning. It outlined the story of a teen named Taylor whose arm was injured playing soccer. The video highlighted the process of diagnosing a fractured arm and covered important information about managing a fractured bone that does not require surgery, including what to expect in the emergency department, cast application and cast care. The video also outlined potential complications and the symptoms requiring immediate care. The video included how to manage pain using both pharmacological and non-pharmacological methods. The video provided concrete examples of how wearing a cast could affect the child’s function including school, sleep and activities of daily living such as bathing, and provided suggestions to lessen this impact. Expected healing timeline and return to normal activities after cast removal was included. Screen captures of the video are included in **Appendix B**.

### Infographic

The interactive infographic looks similar to a webpage and allows users to scroll through the information, exploring at their own pace. The ability for parents to control what they view based on their needs differentiates the interactive infographic from the video. The information provided in the infographic mirrors the information provided in the video and is comprised of 8 major sections: 1) General Information about Fractures, 2) Symptoms, 3) Diagnosis, 4) Casts, 5) Follow-up, 6) Recovery, 7) Recovery Tips, 8) Useful links. Within the follow up section, the infographic provides information on when to seek emergency care including symptoms of serious complications, and when to visit a doctor for less serious potential complications. Within the recovery tips section are several sub-sections outlining helpful information on cast care, sleep, pain management, and helping the child cope. Screen captures of the infographic are included in **Appendix C**.

### Surveys

Parents were recruited to participate in an electronic, usability survey (**Appendix D**) at the Stollery Children’s Hospital emergency department (ED) waiting room in Edmonton. A member of the study team approached parents in the ED to determine interest and study eligibility. Participants who agreed to participate in the study were given an iPad to view the tool and provide their feedback. Surveys comprised of 9, 5-point Likert items (from strongly disagree to strongly agree) assessing usefulness, aesthetics, length, relevance, and future use. Parents were also asked to provide their positive and negative opinions of the tool via two free text boxes. Parents also answered a set of demographic questions.

## Data Analysis

Data was cleaned and analyzed using SPSS v.28. Descriptive statistics and measures of central tendency were generated for demographic questions. Likert scale responses were given a corresponding numerical score, with 5 being “Strongly Agree” and 1 being “Strongly Disagree” [8, 9]. Likert responses were analyzed using means and standard deviations (SD). T-tests were conducted to determine whether there were any significant differences between usability means for the two tools. Open-ended survey data were analyzed thematically. A summary of the results was then shared with the creative team to inform the development of the final versions of the tools.

## Results

Usability of the infographic was evaluated using 9 questions and 2 free text boxes. Each question was rated from *strongly disagree* (1) to *strongly agree* (5). A total of 61 parents awaiting pediatric ED care participated in the study. Of those, 30 viewed the animated video and 31 viewed the interactive infographic. Demographics are shown in **Table 1**.

**Table 1.**
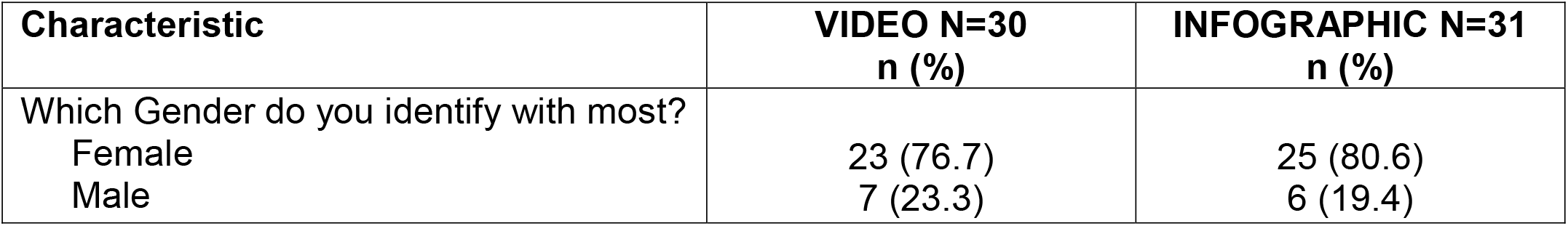
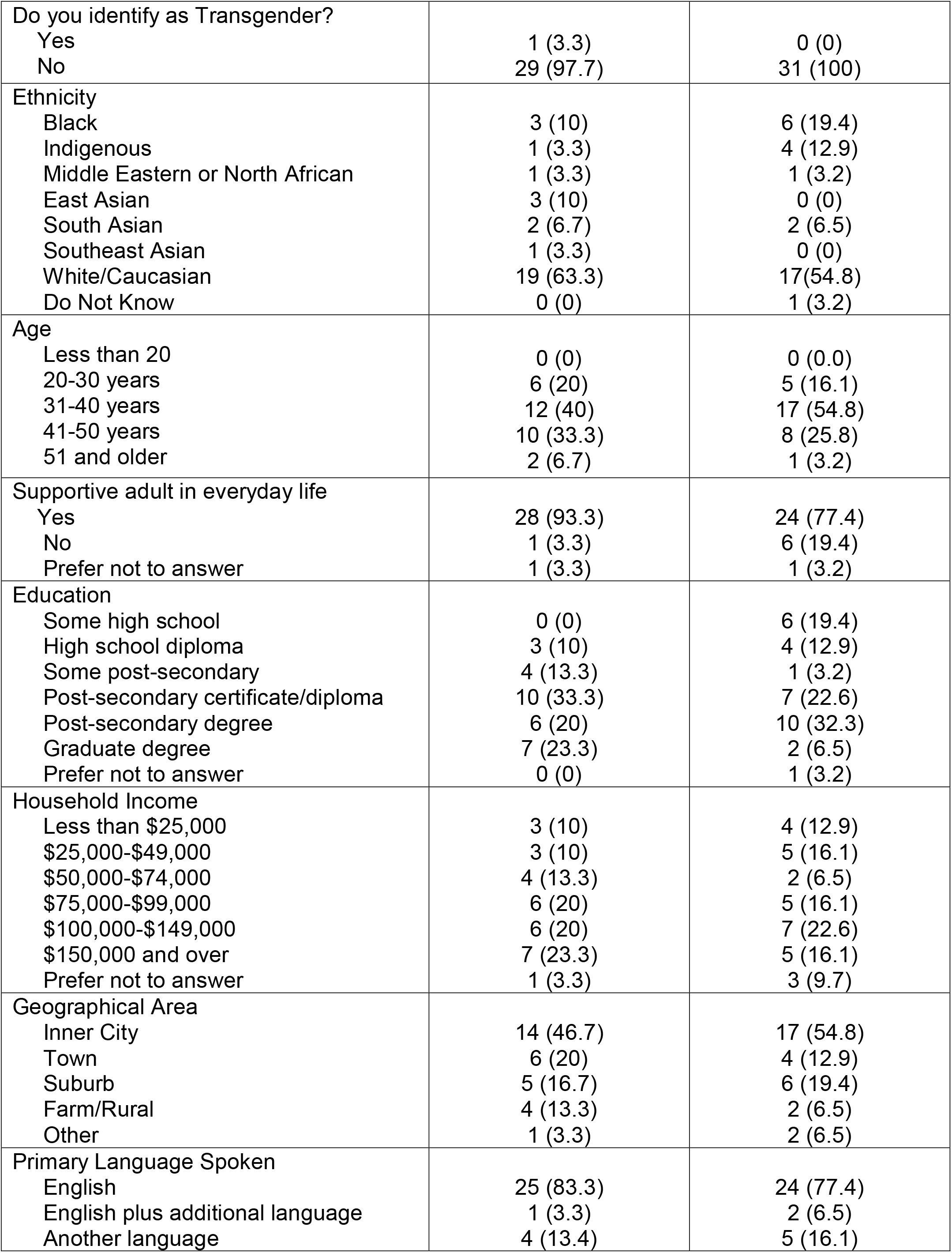
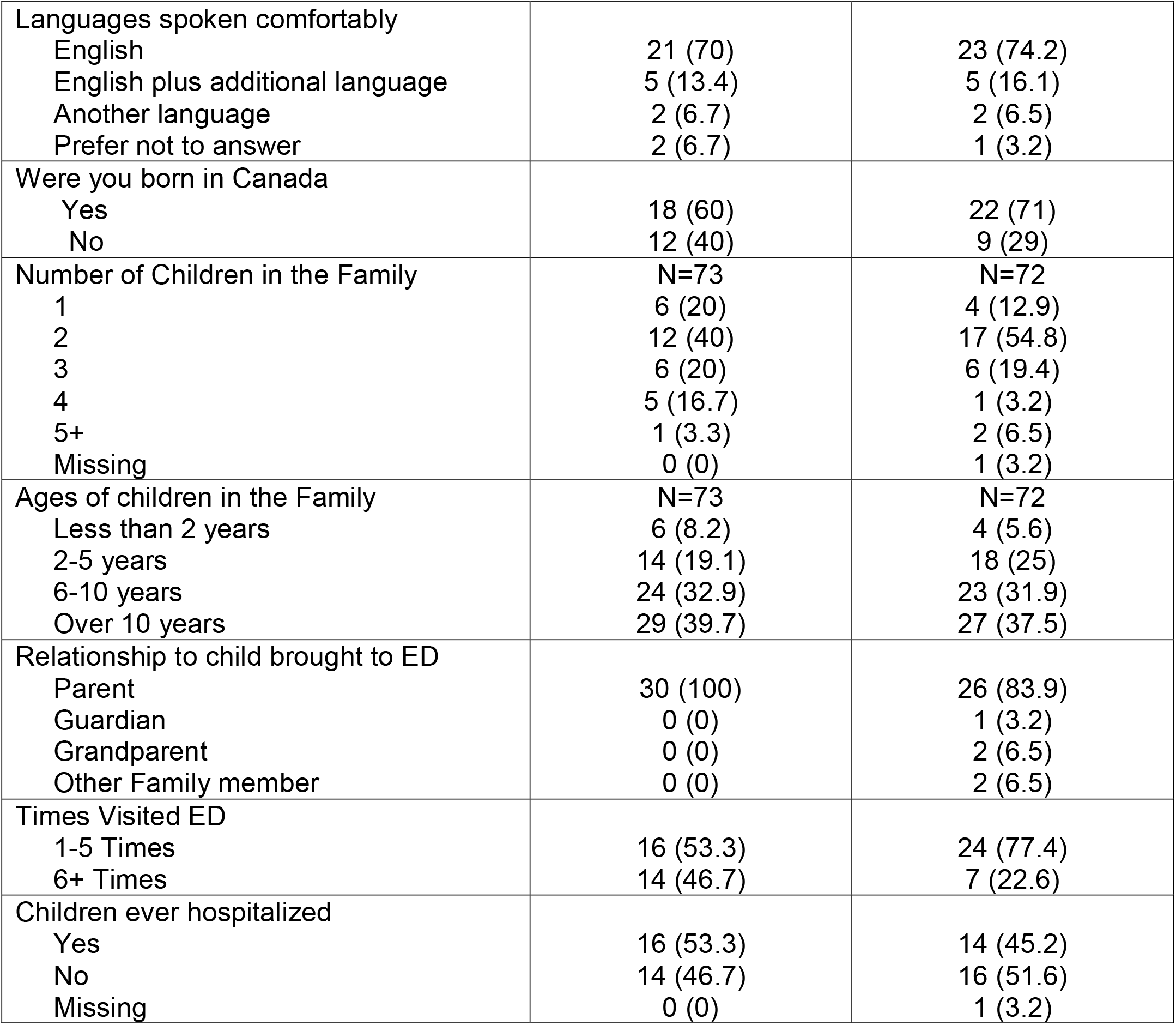
Demographic characteristics of participants who assessed the usability of the video and the infographic.

| Characteristic | VIDEO N=30<br>n (%) | INFOGRAPHIC N=31<br>n (%) |
| --- | --- | --- |
| Which Gender do you identify with most? |  |  |
| Female | 23 (76.7) | 25 (80.6) |
| Male | 7 (23.3) | 6 (19.4) |
| Do you identify as Transgender? |  |  |
| Yes | 1 (3.3) | 0 (0) |
| No | 29 (97.7) | 31 (100) |
| Ethnicity |  |  |
| Black | 3 (10) | 6 (19.4) |
| Indigenous | 1 (3.3) | 4 (12.9) |
| Middle Eastern or North African | 1 (3.3) | 1 (3.2) |
| East Asian | 3 (10) | 0 (0) |
| South Asian | 2 (6.7) | 2 (6.5) |
| Southeast Asian | 1 (3.3) | 0 (0) |
| White/Caucasian | 19 (63.3) | 17 (54.8) |
| Do Not Know | 0 (0) | 1 (3.2) |
| Age |  |  |
| Less than 20 | 0 (0) | 0 (0.0) |
| 20-30 years | 6 (20) | 5 (16.1) |
| 31-40 years | 12 (40) | 17 (54.8) |
| 41-50 years | 10 (33.3) | 8 (25.8) |
| 51 and older | 2 (6.7) | 1 (3.2) |
| Supportive adult in everyday life |  |  |
| Yes | 28 (93.3) | 24 (77.4) |
| No | 1 (3.3) | 6 (19.4) |
| Prefer not to answer | 1 (3.3) | 1 (3.2) |
| Education |  |  |
| Some high school | 0 (0) | 6 (19.4) |
| High school diploma | 3 (10) | 4 (12.9) |
| Some post-secondary | 4 (13.3) | 1 (3.2) |
| Post-secondary certificate/diploma | 10 (33.3) | 7 (22.6) |
| Post-secondary degree | 6 (20) | 10 (32.3) |
| Graduate degree | 7 (23.3) | 2 (6.5) |
| Prefer not to answer | 0 (0) | 1 (3.2) |
| Household Income |  |  |
| Less than \$25,000 | 3 (10) | 4 (12.9) |
| \$25,000-\$49,000 | 3 (10) | 5 (16.1) |
| \$50,000-\$74,000 | 4 (13.3) | 2 (6.5) |
| \$75,000-\$99,000 | 6 (20) | 5 (16.1) |
| \$100,000-\$149,000 | 6 (20) | 7 (22.6) |
| \$150,000 and over | 7 (23.3) | 5 (16.1) |
| Prefer not to answer | 1 (3.3) | 3 (9.7) |
| Geographical Area |  |  |
| Inner City | 14 (46.7) | 17 (54.8) |
| Town | 6 (20) | 4 (12.9) |
| Suburb | 5 (16.7) | 6 (19.4) |
| Farm/Rural | 4 (13.3) | 2 (6.5) |
| Other | 1 (3.3) | 2 (6.5) |
| Primary Language Spoken |  |  |
| English | 25 (83.3) | 24 (77.4) |
| English plus additional language | 1 (3.3) | 2 (6.5) |
| Another language | 4 (13.4) | 5 (16.1) |
| Languages spoken comfortably |  |  |
| English | 21 (70) | 23 (74.2) |
| English plus additional language | 5 (13.4) | 5 (16.1) |
| Another language | 2 (6.7) | 2 (6.5) |
| Prefer not to answer | 2 (6.7) | 1 (3.2) |
| Were you born in Canada |  |  |
| Yes | 18 (60) | 22 (71) |
| No | 12 (40) | 9 (29) |
| Number of Children in the Family | N=73 | N=72 |
| 1 | 6 (20) | 4 (12.9) |
| 2 | 12 (40) | 17 (54.8) |
| 3 | 6 (20) | 6 (19.4) |
| 4 | 5 (16.7) | 1 (3.2) |
| 5+ | 1 (3.3) | 2 (6.5) |
| Missing | 0 (0) | 1 (3.2) |
| Ages of children in the Family | N=73 | N=72 |
| Less than 2 years | 6 (8.2) | 4 (5.6) |
| 2-5 years | 14 (19.1) | 18 (25) |
| 6-10 years | 24 (32.9) | 23 (31.9) |
| Over 10 years | 29 (39.7) | 27 (37.5) |
| Relationship to child brought to ED |  |  |
| Parent | 30 (100) | 26 (83.9) |
| Guardian | 0 (0) | 1 (3.2) |
| Grandparent | 0 (0) | 2 (6.5) |
| Other Family member | 0 (0) | 2 (6.5) |
| Times Visited ED |  |  |
| 1-5 Times | 16 (53.3) | 24 (77.4) |
| 6+ Times | 14 (46.7) | 7 (22.6) |
| Children ever hospitalized |  |  |
| Yes | 16 (53.3) | 14 (45.2) |
| No | 14 (46.7) | 16 (51.6) |
| Missing | 0 (0) | 1 (3.2) |

Participant reactions to the video and infographic were very positive, with all measures obtaining mean results between 4 (agree) and 5 (strongly agree). There were no significant differences between the tools. **Table 2** displays the mean responses to each usability measure for each tool. Comments from the two open-text questions at the end of the usability survey were constructive and mainly positive.

**Table 2.**
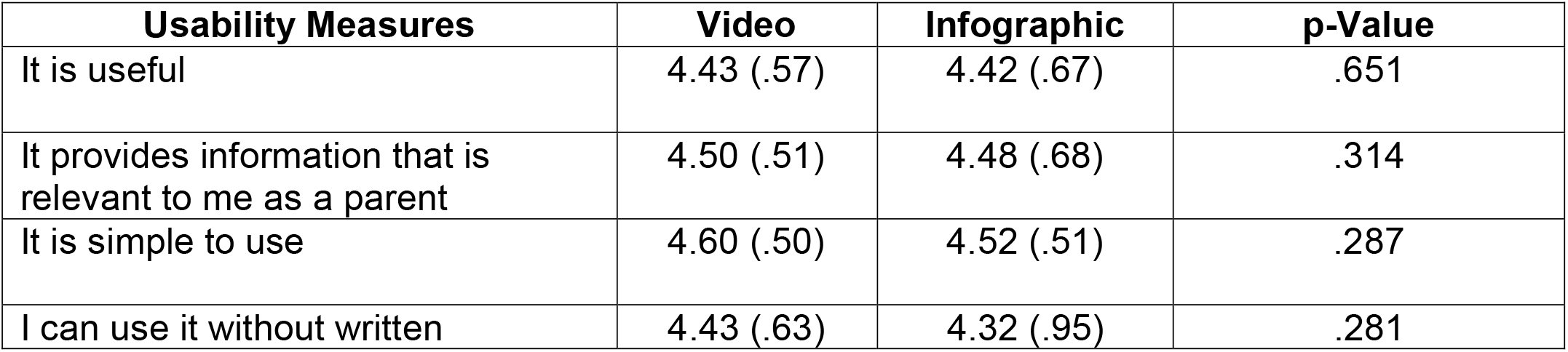
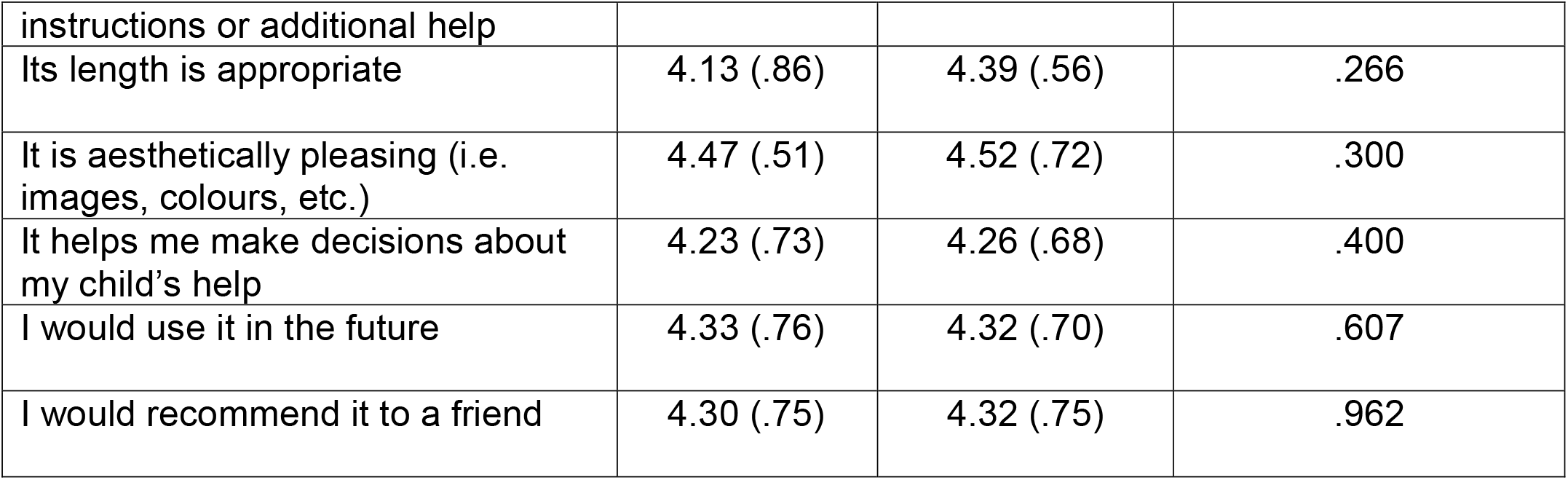
Means (SD) of participant responses to the usability survey.

## Conclusions

Though treatment of simple fractures is relatively straightforward and usually does not require surgery, the recovery process can be long and arduous. The impact of pediatric fractures on the child and their family involves complications such as troubles sleeping, inability to perform daily tasks or take part in activities and play, and changes to daily routines to accommodate functional limitations.

Working with parents, clinical experts, and researchers, we developed two KT tools to help provide parents and children an understanding of the journey to recovery following a pediatric fracture. In collaborating with parents and healthcare providers, we have ensured that the tools are easy to understand, contain relevant information raised by families navigating pediatric fractures, are clinically accurate, and reflect the best evidence. Our tools were highly rated amongst parents seeking care for their children in an urban emergency care centre. Parents found the tools to be useful, relevant, easy to use, and aesthetically pleasing. Most importantly, parents felt that the tools could facilitate decision-making in the future, with parents agreeing and strongly agreeing that they would use the tool and recommend the tool to their friends.

**The tools can be found here: https://www.echokt.ca/fracture/**

Note: Our KT tools are assessed for alignment with current, best-available evidence every two years. If recommendations have changed, appropriate modifications are made to our tools to ensure that they are up-to-date [10]. The fracture KT tools are up to date as of April 2025.

## Data Availability

All data produced in the present study are available upon reasonable request to the authors.

## Author Contributions

This research was conducted under the supervision of Dr. Shannon D. Scott (SDS), PI for **translation Evidence in Child Health to enhance Outcomes** (ECHO) Research, Dr. Lisa Hartling (LH), PI for the **Alberta Research Centre for Health Evidence** (ARCHE), Dr. Samina Ali (SA), PI for the **Pediatric Emergency Advancing Knowledge** (PEAK) Research Team, and Dr. Manisha Bharadia (MB). SDS and LH designed the research study and obtained research funding through the Stollery Science Lab Distinguished Researchers program which is funded by the Stollery Children’s Hospital Foundation and the Women and Children’s Health Research Institute.

SDS designed and supervised all aspects of tool development and evaluation. LH contributed to tool development and evaluation. SA and MB conducted and analyzed qualitative interviews with parents and adolescents in previous studies (1, 2) which informed this work.

SA, MB, and Hannah M. Brooks (HMB) contributed to tool development and evaluation. HMB conducted usability testing and analyzed usability data.

All authors contributed to the writing of this technical report and provided substantial feedback.

**This work was funded by:**

**Women and Children’s Health Research Institute and the Stollery Children’s Hospital Foundation:**

• **Scott, S.D**., Hartling, L. Distinguished Researcher Funding. (2018). Women and Children’s Health Research Institute (WCHRI) & Stollery Children’s Hospital Foundation ($1,000,000). September 2018-January 2025.

**This report should be cited as:** Scott, S.D., Brooks, H.M., Ali, S., Bharadia, M., Hartling, L. (2026). The development and usability testing of two arts-based knowledge translation tools for pediatric fractures. Internal Technical Report. ECHO Research, University of Alberta.

*Available at*: http://www.echokt.ca/research/technical-reports/

## Appendices

## Appendix A

**Video Screen Captures**

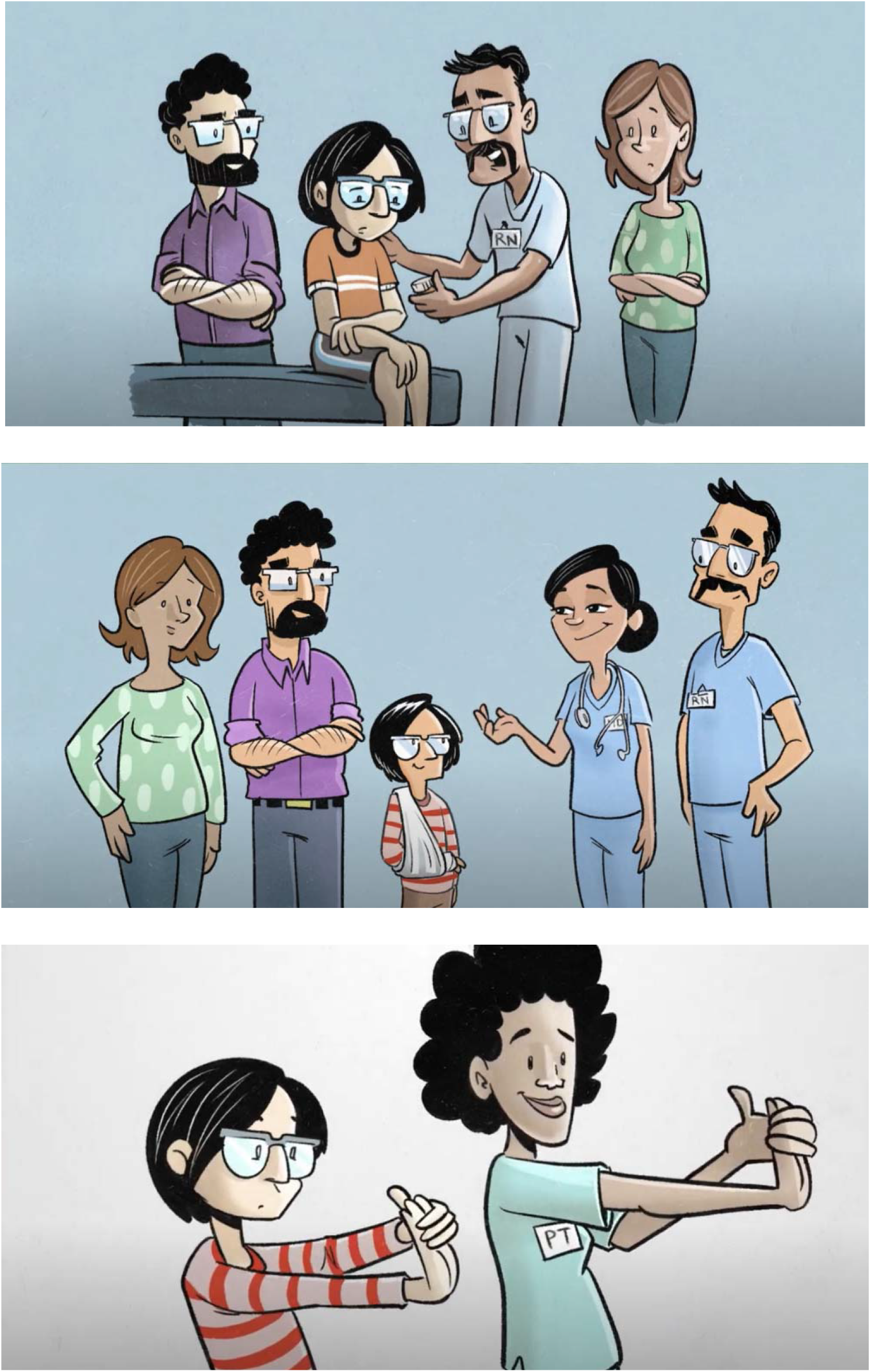

## Appendix B

**Infographic Screen Captures**

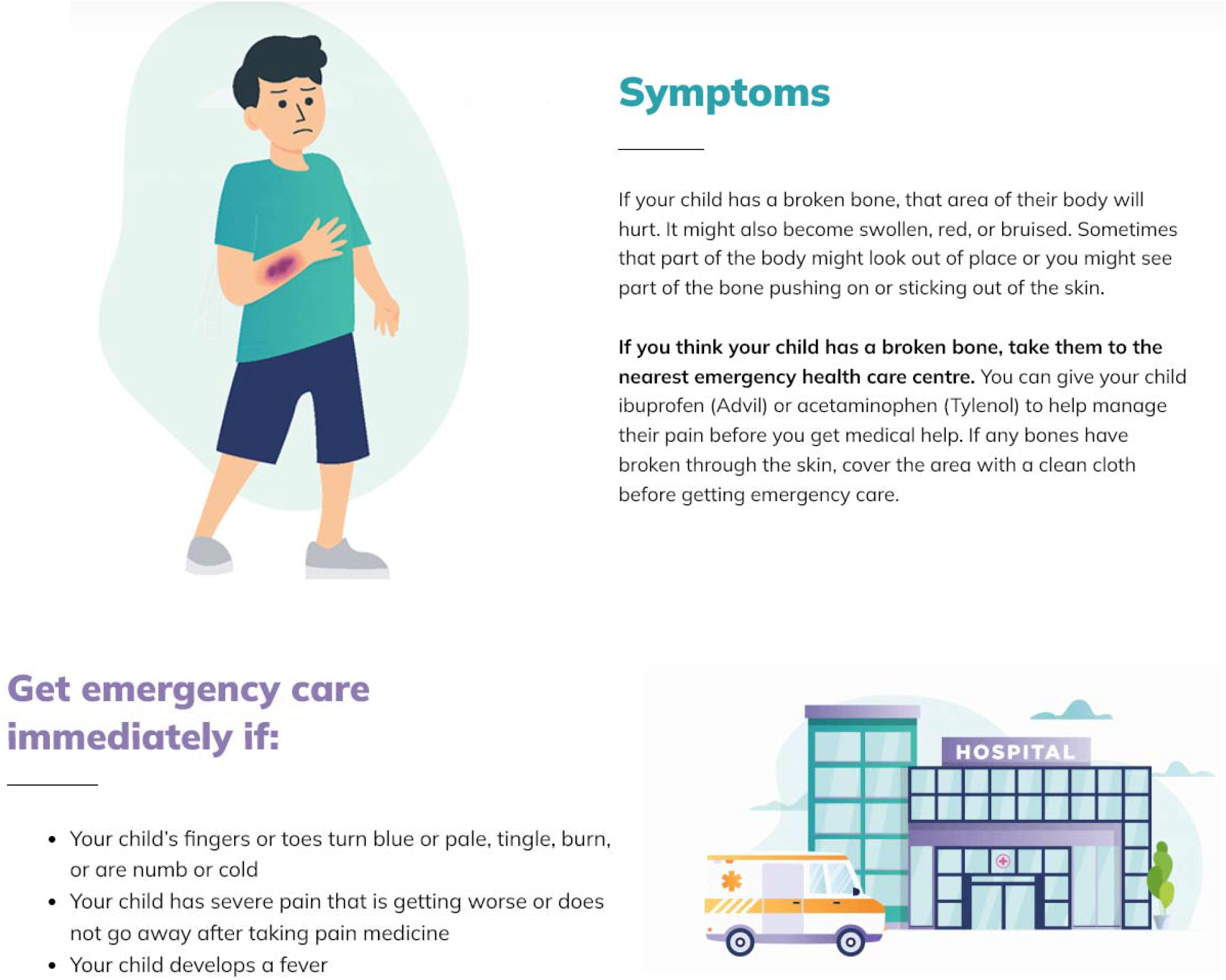

## Appendix C

**Usability Survey**

SECTION 1: Demographics

1) a. What is your gender?
  □ Male
  □ Female
  □ Non-binary
  □ Gender fluid
  □ Not sure or questioning
  □ Another referred term ___________
  □ Prefer not to answer

1. b. Would you describe yourself as transgender?
  □ Yes
  □ No
  □ Prefer not to answer
2. Which ethnicities best describes you? *Please select all that apply*.
  □ Black (includes African, Afro-Caribbean, African Canadian descent)
  □ East Asian (includes Chinese, Korean, Japanese, Taiwanese descent)
  □ Indigenous (includes First nations. Metis, Inuk/Inuit descent)
  □ Latino (includes Latin American, Hispanic descent)
  □ Middle Eastern or North African (includes Arab, Persian, Afghan, Egyptian, Iranian, Lebanese, Turkish, Kurdish and other West Asian descent)
  □ South Asian (includes East Indian, Pakistani, Bangladeshi, Sri Lankan, Indo-Caribbean, and other South Asian descent)
  □ Southeast Asian (includes Filipino, Vietnamese, Cambo
dian, Thai, Indonesian and other SouthEast Asian descent)
  □ White (includes European descent)
  □ Not listed or other: ___________
  □ Do not know
  □ Prefer not to answer
3. What is your age?
  □ Less than 20 years old
  □ 20-30 years
  □ 31-40 years
  □ 41-50 years
  □ 51 years and older
4. Do you have a supportive adult in your everyday life?(e.g. spouse, common-law partner, other partner/relationship)
  □ Yes
  □ No
  □ Prefer not to answer
5. What is your gross annual household income?
  □ Less than $25,000
  □ □ $25,000-$49,999
  □ □ $50,000-$74,999
  □ □ $75,000-$99,999
  □ □ $100,000-$149,999
  □ □ $150,000 and over
  □ Prefer not to answer
□ What is your highest level of education?
  Some high school
  □ High school diploma
  □ Some post-secondary
  □ Post-secondary certificate/diploma
  □ Post-secondary degree
  □ Graduate degree
  □ Other: ___________
  □ Prefer not to answer
7. Where does your household live
  □ Inner City
  □ Suburb
  □ Town
  □ Farm/Rural
  □ Other: ___________
8. How many children do you have?_______
9. What are the ages are your children?_______
10. What is the primary language spoken in your home?_______
11. What languages can you speak comfortably?_______
12. Were you born in Canada?
  □ Yes
  □ No
  □ Prefer not to answer
13. What is your relationship to the child that you have brought to the emergency department?
  □ Parent
  □ Grandparent
  □ Other family member
  □ Guardian
14. How many times have you visited the emergency department with your children?
  □ 1-5 times □ 6+times
□ Have any of your children ever been admitted to the hospital?
  □ Yes
  □ No

SECTION 2: Assessment of attributes of the arts-based, digital tools.

*Items 1 to 9 were assessed using a 5-point Likert Scale where 1=strongly disagree and 5=strongly agree*.

1. It is useful.
2. It provides information that is relevant to me as a parent.
3. It is simple to use.
4. I can use it without written instructions or additional help.
5. Its length is appropriate.
6. It is aesthetically pleasing (i.e., images, colours, etc.).
7. It helps me to make decisions about my child’s health.
8. I would use it in the future.
9. I would recommend it to a friend.
10. List the most positive aspects: [open text]
11. List the most negative aspects: [open text]

## Appendix D

**Project Timeline**

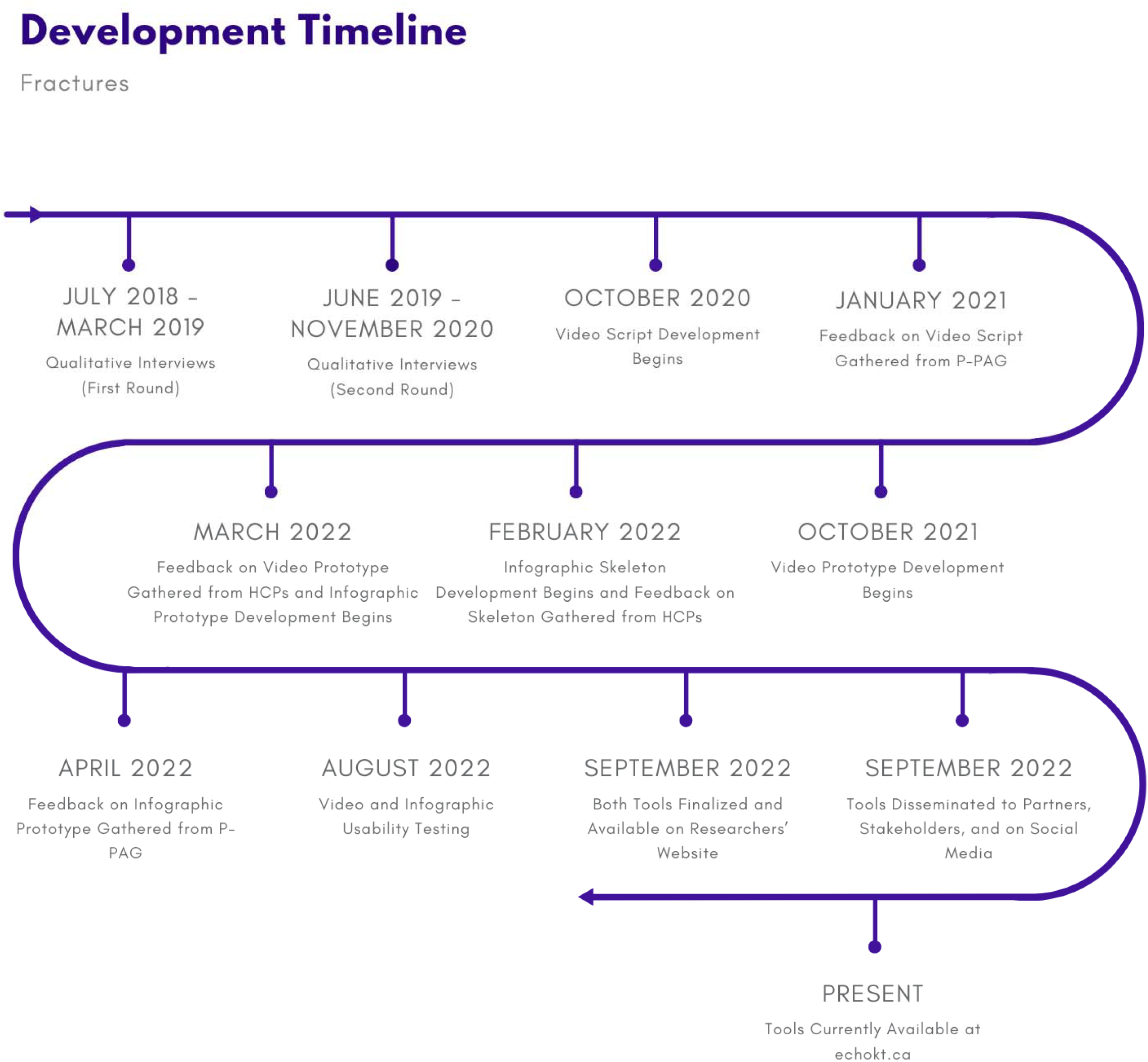

HCPs, healthcare providers; P-PAG, Pediatric-Parents’ Advisory Group.

